# A systematic review to assess seizure risk with chloroquine therapy in persons with epilepsy

**DOI:** 10.1101/2020.04.09.20056358

**Authors:** Sandipan Pati

## Abstract

**Background:** The goal of this systematic review is to assess the published literature for seizure risk with chloroquine therapy in persons with and without epilepsy. With the COVID-19 pandemic, there is a desperate need for therapy against the SARS CoV-2 virus. Chloroquine is one proposed medication that has received substantial public attention. However, drug labeling in the package insertion states that persons with epilepsy have the risk of chloroquine provoking seizures, and this has increased questions and anxiety in the epilepsy community.

**Methods:** PubMed (1970 to March 27, 2020) and the Embase (1970 to March 27, 2020) were searched with the terms chloroquine and seizure or epilepsy. Selected studies were reviewed, and the adverse drug reaction was classified.

**Results:** Only nine out of 27 studies were deemed eligible for systematic analysis. Out of the nine studies, only one was a prospective study (N=109), two were case series (N=6), and the remaining 6 were case reports. The dose of chloroquine ranged between 100-500 mg/day, except in one patient, the seizure was after taking 1000 mg. The strength of causality for the drug causing seizures in healthy and persons with epilepsy was mostly possible or unlikely, and none were certain. The only clinical trial that evaluated seizure risk with chloroquine failed to find any significant relation.

**Conclusion:** Although the drug insertion label states an increased risk of seizure, the systematic review highlights that such a statement is not supported by any class I studies but by anecdotal case reports. The only randomized clinical study revealed that seizures were not associated with an increased blood level of chloroquine or its metabolite. The present systematic review should provide reassurance to busy clinicians and persons with epilepsy that chloroquine, if prescribed to treat COVID-19, lacks any substantial evidence to suggest that the medication increases the risk of seizure.

## INTRODUCTION

With the global outbreak of coronavirus disease (COVID-19), laboratory-confirmed cases have increased explosively, with 68,000 died as of April 4, 2020(WHO, 2020). A significant proportion of symptomatic patients required medical care and some required specialized intensive care management that is overwhelming the health care system. Given the acute shortage of ventilators and personal protective equipment, there is a desperate need for therapy against the virus SARS CoV-2 - the pathogen for COVID-19(Fauci et al., 2020). Among the different medications that are under trial, two antimalarial drugs-chloroquine and hydroxychloroquine have received substantial public attention because of positive results from small studies and media coverage(Cortegiani et al., 2020)(Hu et al., 2020). A quick search in the clinicaltrials.gov with the term chloroquine or hydroxychloroquine for COVID-19 yielded over 30 studies as of early April 2020. Despite the lack of any substantial evidence to support efficacy, given the present crisis, many academic centers and physicians have incorporated these medications in their therapeutic armamentarium against COVID-19(Cortegiani et al., 2020). The drug labeling in the package insertion for chloroquine states that “patients with a history of epilepsy should be advised about the risk of chloroquine provoking seizures.” Understandably this statement has stirred up increased questions and anxiety in the epilepsy community about the safety of chloroquine in persons with epilepsy. The goal of this systematic review is to assess the published literature for seizure risk with chloroquine therapy in persons with and without epilepsy.

## METHODS

This study was performed in accordance with the recommendations of the Preferred Reporting Items for Systematic Reviews and Meta-analyses (PRISMA) reporting guidelines. PubMed (1970 to March 27, 2020) and the Embase (1970 to March 27, 2020) were searched with the terms chloroquine and seizure or epilepsy. Only published abstracts in English were reviewed. Data extracted from the reports were -age, sex of study populations, the dosage of medication if available, and reported comorbidities. Exclusion criteria were overdose or poisoning from chloroquine, animal studies, and reports of cardiac or neuropsychiatric adverse effects.

The adverse drug reaction of interest is a seizure or status epilepticus. For each selected study, the author reviewed the entire manuscript and classified the adverse drug reaction as either– a) dose-related, b) non-dose related, c) dose-related and time-related, d)time-related, e) withdrawal or f) unexpected failure of therapy(Edwards and Aronson, 2000). Finally, the author classified the causality of the drug (i.e., the chloroquine) inducing adverse drug reaction (i.e., the seizure) as – a) certain, b) probable or likely, c) possible, d) unlikely, e) conditional, or unclassified and f) unassessable or unclassifiable(Edwards and Aronson, 2000).

## RESULTS

Only nine out of 27 studies were deemed eligible for systematic analysis. The excluded studies were either preclinical or reports of chloroquine poisoning or cardiovascular and neuropsychiatric complications. Out of the nine studies, only one was a prospective study(N=109), two were case series(N=6), and the remaining 6 were case reports (Table 1).

**Table 1:** Characteristics of nine studies (six case reports, two case series, and one clinical trial)

| Type of Adverse Reaction | Type of study | Patient comorbidities | Drug/dose | Reason for prescription | Seizures or status epilepticus (SE) | Type of association | Country reported | References |
| --- | --- | --- | --- | --- | --- | --- | --- | --- |
| Non dose related | Case report | 49 yo, F, SLE | C , 250 mg/dX 30 d | SLE | multiple FIAS X 24 hrs | Possible (CSF not analyzed, EEG abnormal) | Poland | 2018. Krzeminski P, Lesiak A, Narbutt J. PMID: 30206460 |
| Non dose related | Case report | 14 yo, F, SLE | C, 500mg/d | SLE | one BTC sz | Probable (CSF negative, EEG normal) | Venezuela | 2004 Tristano AG, Falcón L, Willson M, de Oca IM PMID: 14740170 |
| dose related | Case report | 30 yo,M, healthy | C, 1gm/d X4 d | Ppx | one BTC sz | Possible (CSF, EEG not performed) | UK | 2016 Martin AN, Tsekas D, White WJ, Rossouw D. PMID: 27005796 |
| Non dose related | Case report | 35 yo,M, healthy | Savarine (C 100 mg+Proguanil 200 mg) -2 doses | Ppx | one BTC sz | Possible (CSF, EEG not performed) | France | 2000. Schiemann R, Coulaud JP, Bouchaud O. PMID: 11179947. |
| Non dose related | Case series | 40 yo M, healthy | Maloprim (C 400 mg+ Dapsone 100 mg+ Pyrimthimine 12.5 mg) 1 tab/wk X 4 wks | Ppx | two BTC sz | Unlikely (CSF not analyzed, EEG abnormal-generalized spike wave) | UK | 1988, Fish DR, Espir ML. PMID: 3139186 |
|  |  | 26 yo F, generalized epilepsy | Maloprim (C 400 mg+ Dapsone 100 mg+ Pyrimthimine 12.5 mg) 1 tab/wk X 3 wks | PPx | one BTC sz | Unlikely(CSF not analyzed, EEG abnormal) |  |  |
|  |  | 49 yo F, focal epilepsy | C 400 mg /d X 1 d | Ppx | prolonged BTC sz | Unlikely (CSF not analyzed, EEG abnormal) |  |  |
| Non dose related | Case report | 42 yo, M, Leprosy | C 450 mg/d x 8 d | ENL (Leprosy) | two BTC szs | Possible(CSF negative, EEG not performed) | Nigeria | 1998 Ebenso BE. PMID: 9715604 |
| Non dose related | Double-blinded randomized clinical trial | N=109; 9 mo-13 y, cerebral malaria | dose unknown, study correlated seizure with serum level of C and a metabolite | Ppx | 54% szs, 9% SE | Unlikely ( no coorelation between serum level and seizure) | Kenya | 2000. Crawley J, Kokwaro G, Ouma D, Watkins W, Marsh K. PMID: 11169275 |
| Non dose related | Case series | 28 yo, F, SLE | C 200 mg/d X 180d | SLE | one BTC sz | Probable(CSF negative, EEG post ictal slowing) | Netherlands | 1992. Luijckx GJ, De Krom MC, Takx-Kohlen BC. PMID: 1344765 |
|  |  | 21,yo, M, healthy | C 300 mg /week X 2mo | Ppx | two BTC sz | Probable (CSF negative, EEG normal) |  |  |
|  |  | 23 yo, M, h/o one sz | C 300 mg /week X 2mo | Ppx | one BTC sz | Possible (CSF not analyzed, EEG normal) |  |  |
| Non dose related | Case report | 69=8 yo, F, healthy | C 100mg/d X12 d | Ppx | NCSE | Probable (CSF negative, EEG showed SE) |  | 1995. Mülhauser P, Allemann Y, Regamey C.PMID: 7762925 |

F= female, M=male, C=chloroquine, Ppx= prophylaxis, SLE= systemic lupus erythematosus, ENL= erythema nodosum leprosum, FIAS= focal impaired awareness seizure, BTC= bilateral tonic-clonic seizure, CSF= cerebrospinal fluid

### a) Risk in healthy individuals

Pooled data revealed five healthy adults (all case reports and series) had a convulsive seizure after taking chloroquine for primary prophylaxis against malaria. One of the individuals had speech difficulties secondary to non-convulsive status epilepticus. The dose ranged between 100 mg to 400 mg per day except in an individual who took 1 gm that was prescribed by a non-professional personal. The causality between the seizure and the chloroquine is determined to be “possible” for two of them as alternative causes of seizures were not explored while in two of them as “probable” after extensive investigations ruled out alternative etiology. In one individual, EEG revealed generalized spike-wave epileptiform discharges, thereby suggesting underlying undiagnosed generalized epilepsy.

### b) Risk in individuals with non-neurological illness

Four adults (all case reports and series) had a convulsive seizure after taking therapeutic chloroquine for leprosy or lupus. The dose ranged between 200-500 mg/day. The causality between the seizure and the chloroquine is determined to be “probable” for two of them after multiple laboratory investigations, including CSF analyses, and EEG failed to identify any alternative etiology.

### c) Risk in individuals with neurological illness, including epilepsy

One double-blinded, randomized clinical trial involving 109 children (9 mo-13 years) with cerebral malaria failed to find any correlation between seizure and blood level of chloroquine or desethylchloroquine-a metabolite of chloroquine(Crawley et al., 2000). This is the only clinical trial that evaluated seizure risk with chloroquine and found no significant relation(Crawley et al., 2000). Three case studies documented seizure following chloroquine prophylaxis(300-400 mg/day) in persons with epilepsy. None of the associations were probable, and at best, were possible.

## DISCUSSION AND CONCLUSION

The sulfate and phosphate salts of chloroquine have been used as antimalarial drugs and in treating lupus over a decade. Although the drug insertion label states an increased risk of seizure, the systematic review highlights that such a statement is not supported by any class I studies but by anecdotal case reports. The only randomized clinical study revealed that seizures were not associated with an increased blood level of chloroquine or its metabolite. None of the case reports or series proved the causality of chloroquine provoking seizures to be certain.

The goal of this review was not to evaluate if chloroquine is an effective therapy against COVID-19. Clinical trials are ongoing, and many centers are offering the medication either through a clinical trial or as an off label therapy. Many of our epilepsy patients reside at a group home or facility and are at risk of contracting SARS CoV-2 virus. The present systematic review should provide reassurance to busy clinicians and persons with epilepsy that chloroquine, if prescribed to treat COVID-19, lacks any substantial evidence to suggest that the medication increases the risk of seizure.

## Data Availability

this is no clinical data. This is a systematic review

